# Acute Shortage Ventilator

**DOI:** 10.1101/2020.07.20.20158147

**Authors:** D. S. Akerib, A. Ames, M. Breidenbach, M. Bressack, P. A. Breur, E. Charles, D. Gaba, R. Herbst, C. M. Ignarra, S. Luitz, E. H. Miller, B. Mong, T. A. Shutt, M. Wittgen

## Abstract

We have implemented an “Acute Shortage Ventilator” (ASV) motivated by the COVID-19 pandemic and the possibility of severe ventilator shortages in the near future. The unit cost per ventilator is less than $400 US excluding the patient circuit parts. The ASV mechanically compresses a self-inflating bag resuscitator, uses a modified patient circuit, and is commanded by a microcontroller and an optional laptop. It operates in both Volume-Controlled Assist-Control mode and a Pressure-Controlled Assist-Control mode. It has been tested using an artificial lung against the EURS guidelines. The key design goals were to develop a simple device with high performance for short-term use, made primarily from common hospital parts and generally-available non-medical components, and at low cost and ease in manufacturing.

## 1 Introduction

We have implemented an “Acute Shortage Ventilator” (ASV) motivated by the COVID-19 pandemic and the possibility of severe ventilator shortages in the near future. This ventilator is based on a simple mechanical design, powered by compressed air to a pneumatic cylinder which compresses a standard self-inflating bag resuscitator. It utilizes a patient circuit made from standard medical components. The pressure in the patient circuit is limited by a Peak Inspiration Pressure (PIP) valve, constructed by a simple modification of a commercial Positive End-Expiratory Pressure (PEEP) valve. The flow and pressure in the patient circuit are monitored by a commercial single-use spirometer feeding a differential pressure sensor. An Arduino single-board microcontroller unit (MCU) reads the sensors and controls the pneumatic valves that operate the ASV. The system can run autonomously using a local display and control knob to adjust parameters. More extensive monitoring of the patient pressure and flow waveforms, and all other system details, can be done using a laptop and the provided Graphical User Interface (GUI).

The intellectual property of the ASV will be held by Stanford University and made available to vendors and manufacturers via no-cost licenses.

## 2 System Requirements

The ASV is designed to meet the Association for the Advancement of Medical Instrumentation (AAMI) Emergency Use Resuscitator System (EURS) design guidance specifications (AAMI/CR503:2020 [4]). These are modifications to the ISO specifications for critical care ventilators (ISO:80601-2-12 [6]) and the general standards for electrical medical equipment (IEC:60601-1 [7]) that are applicable to simplified ventilator designs that consist of mechanical systems to squeeze self-inflating bag resuscitators.

### 2.1 Fundamental Requirements

As summarized in the EURS (e.g., EURS lines, 43-46), ventilator support needs of a COVID-19 patient can include:

1. Bi-level positive airway pressure (BIPAP) for patients breathing spontaneously (EURS-43);
2. Pressure-control mandatory breathing controlled by ventilator (EURS-44);
3. Volume-control mandatory breathing controlled by ventilator (EURS-45);
4. Increased inspired oxygen concentration, or FiO_2_ *>* 21% (Fraction of Inspired Oxygen) (EURS-46).

The ASV operates by mechanically squeezing a self-inflating resuscitator bag to deliver either a controlled tidal volume or a controlled peak inspiratory pressure (PIP). Deliverance of a breath can be initiated by a set respiratory rate or by a spontaneous inhalation from the patient with a set Trigger Threshold. The last operating mode can be achieved with simple modifications to the operation by attaching the bag to a supply of concentrated oxygen. These modes are described in more detail in §3.2.

### 2.2 Requirements on Controllable Parameters, Monitoring and Alarms

The EURS also summarizes sets of parameters that must be controlled and monitored to be able to properly manage a COVID-19 patient (summarized in Table 1).

**Table 1:** Summary of requirements on key control and monitoring parameters. The tolerance on the tidal volume is reported as *±* bias *±* linearity. The narrower tidal volume range is the requirement, the wider range is the recommendation as specified in EURS. The Alarms column lists the alarm condition associated with each parameter, The Reference column gives the line number in the EURS document that describes the requirement on that parameter.

| Parameter | Range | Tolerance | Alarms | Reference |
| --- | --- | --- | --- | --- |
| I:E Ratio | 1:1 – 1:4 | – | – | EURS-56 |
| Respiratory rate (BPM) | 10 – 30 | $\pm 2$ | – | EURS-57 |
| Tidal volume (ml) | 350 – 450 (250 – 600) | $\pm 4\% \pm 15\%$ | $\pm 20\%$ | EURS-59 |
| Inspiratory pressure (cm H<sub>2</sub>O) | < 40 | $\pm 4\%$ | High, Low, Cont. | EUR-256 |
| FiO<sub>2</sub> (%) | External Control |  |  |  |

The control and monitoring of these key parameters as well as a number of derived and related parameters are described in §3.3. Control of the FiO_2_ is established by connecting the ASV to an external oxygen supply. Alarm conditions for these parameters are generated in compliance with clause 12 of the ISO:80601-2-12 specification of the EURS document. They are summarized in Table 2 and further described in §3.4. The ASV has not been submitted to the US FDA for certification.

**Table 2:**
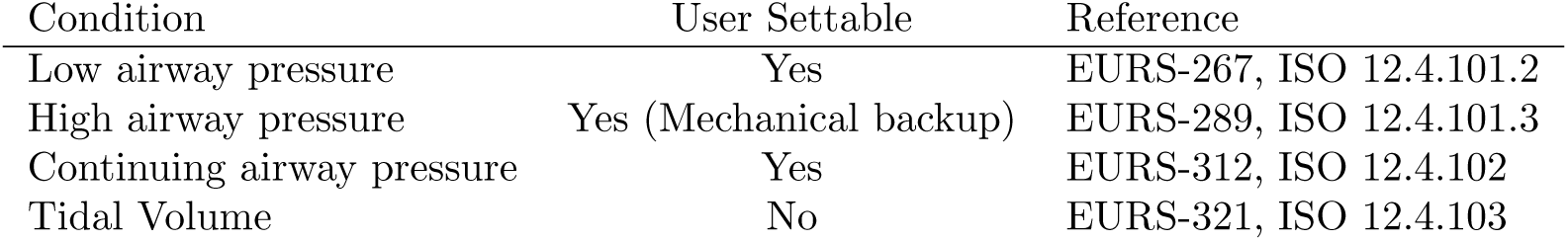
Summary of alarm conditions related to key control parameters. For the settable alarm conditions the alarm limits are user-controllable. The high airway pressure alarm requirement also specifies that there must be a mechanical backup that prevents the airway pressure from exceeding 60 cm H_2_O.

| Condition | User Settable | Reference |
| --- | --- | --- |
| Low airway pressure | Yes | EURS-267, ISO 12.4.101.2 |
| High airway pressure | Yes (Mechanical backup) | EURS-289, ISO 12.4.101.3 |
| Continuing airway pressure | Yes | EURS-312, ISO 12.4.102 |
| Tidal Volume | No | EURS-321, ISO 12.4.103 |

### 2.3 General Requirements for Electrical Medical Systems

The EURS also specifies a set of general safety guidelines, most of which are derived from the general specification for safety of electrical medical devices, IEC:60601-1, (see in particular clauses 5-17 of that document). These cover a number of items including labeling, electrical and mechanical safety, and documentation.

Although certifying that a device meets IEC:60601-1 requirements is an involved process, in the case of the ASV, this will be greatly simplified for a number of reasons. Specifically:

- The EURS specifically excludes some of the requirements, and clarifies relatively simple ways in which certain requirements may be met; e.g., the requirements on a backup system in the case of power failure may be met with the use of an external Uninterruptible Power Supply (UPS).
- The ASV is electrically isolated from the patient by the entire patient breathing circuit.
- Internally, the ASV uses only 12 V power, thus mitigating many potential electrical hazards.
- The ASV electronics are all enclosed.

## 3 Description of System

A schematic overview of the system and a photograph of a prototype are shown in Figure 1. The ASV consists of the patient circuit, the mechanical and pneumatic system that compresses the self-inflating bag, the sensors associated with the spirometer, electronics, and firmware/software.

**Figure 1:**
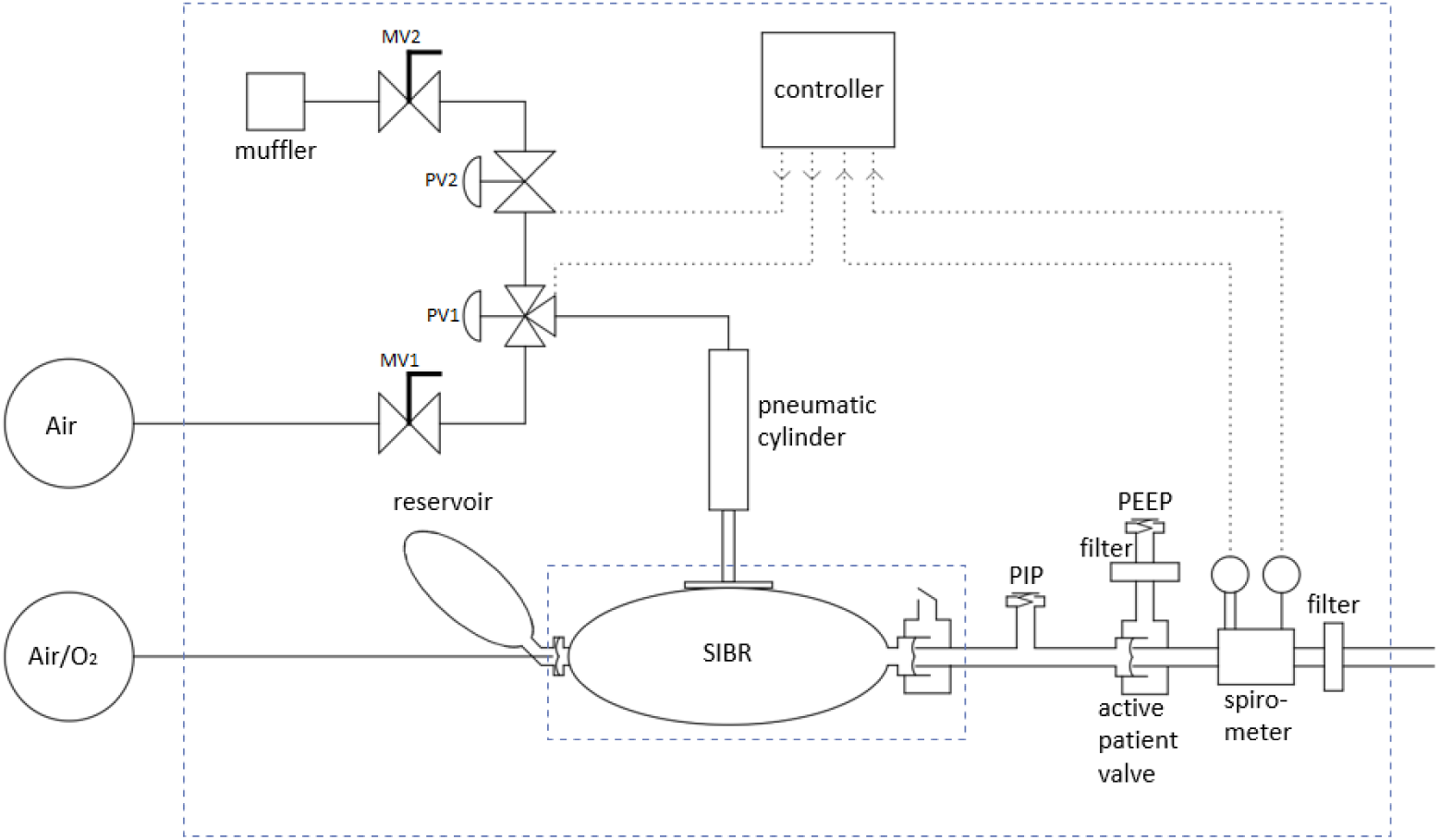
System diagram of the ASV. The SIBR represents a standard unmodified self-inflating bag resuscitator. The patient circuit to the right consists of standard ICU parts except for a modified PEEP Valve, the pressure and flow sensors, and two adapters.

### 3.1 Patient Circuit

The ASV patient circuit is built primarily from standard hospital components. It operates in a volume- or pressure-limited assist control mode. The patient air supply may be ambient room air or an externally supplied mixture of air and oxygen. The self-inflating bag is compressed for each inhalation cycle, and is capable of delivering up to 700–800 ml of the air/oxygen mix. The bag output is pressure-limited by the mechanical PIP Valve. A hose connects the PIP Valve tee to the remaining components near the patient. There, an Active Patient Valve serves as a check valve to minimize dead space and directs the exhalation air stream to a separate port. Next, a spirometer measures flow and pressure, followed by a Heat and Moisture Exchanger and Filter (HMEF), and finally, connects to the endotracheal tube (ETT). A PEEP Valve is added to the exhaust port of the Active Patient Valve, ideally with a HEPA filter to minimize contagion. Positive End-Expiratory Pressure — PEEP — is a setting of a ventilator system that ensures that the pressure at the end of either a mechanical or spontaneous breath remains above ambient. This is done to keep the small air sacs (alveoli) open when they are prone to collapse in different sorts of lung disease, including ARDS. The complete patient circuit is shown in Figure 2.

**Figure 2:**
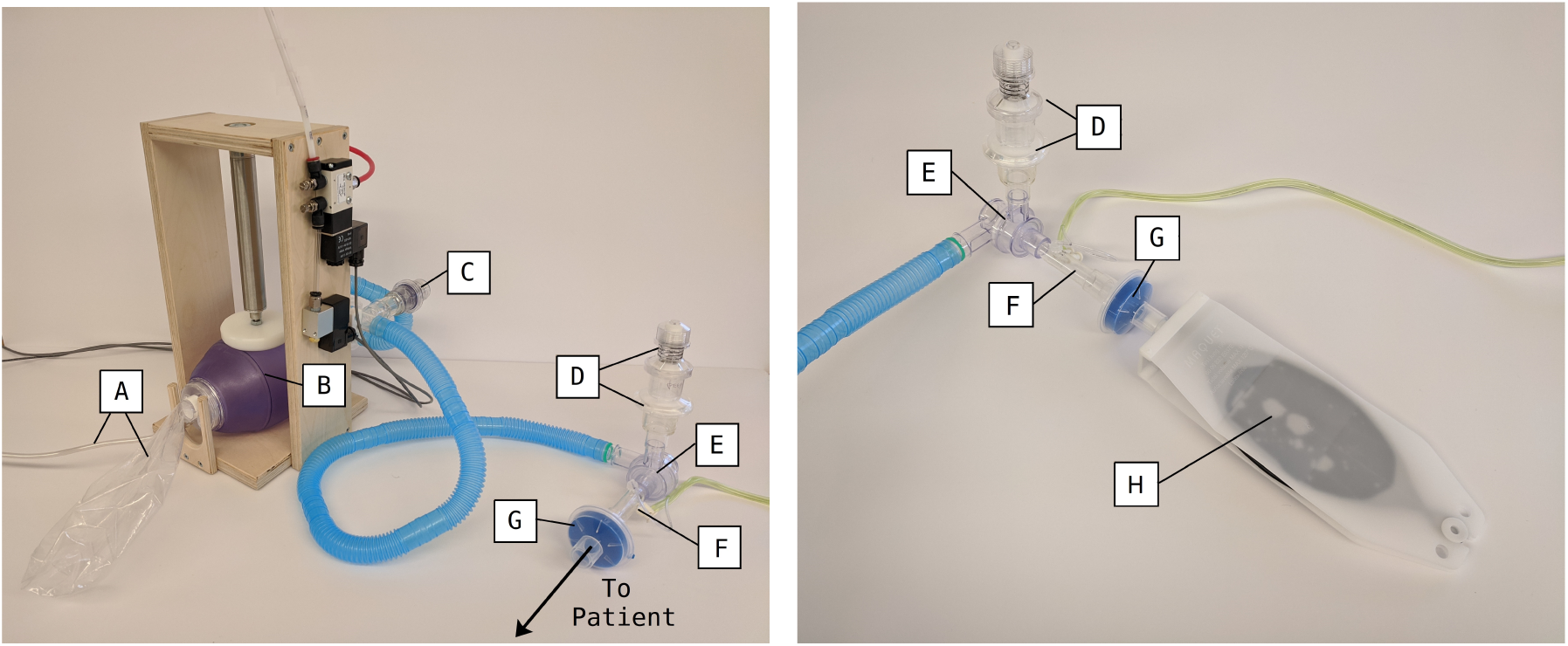
Close up of patient circuit (left) and patient connection to test lung (right). A: Patient air supply hose and reservoir bag; B: Self-inflating bag; C: PIP Valve; D: HEPA filter and PEEP Valve; E: Active patient valve; F: Spirometer; G: HMEF filter; H: Simple test lung.

This entire patient circuit can be assembled from standard, disposable hoses and components routinely used in hospitals, with one modification. The PIP Valve is constructed from a PEEP Valve by inserting a spring-compressing spacer into the valve. This results in a PIP Valve with a pressure range of approximately 20 to 40 cm H_2_O.

### 3.2 Modes of Operation

The ASV GUI is shown in Figure 3. The ASV can operate in Volume Controlled Assist Control (VC-AC) mode or Pressure Controlled Assist Control (PC-AC) mode. In both modes, the Respiration Rate (RR) and the Inspiratory Time can be set, as can the Inspiratory pressure that will trigger an assist breath.

**Figure 3:**
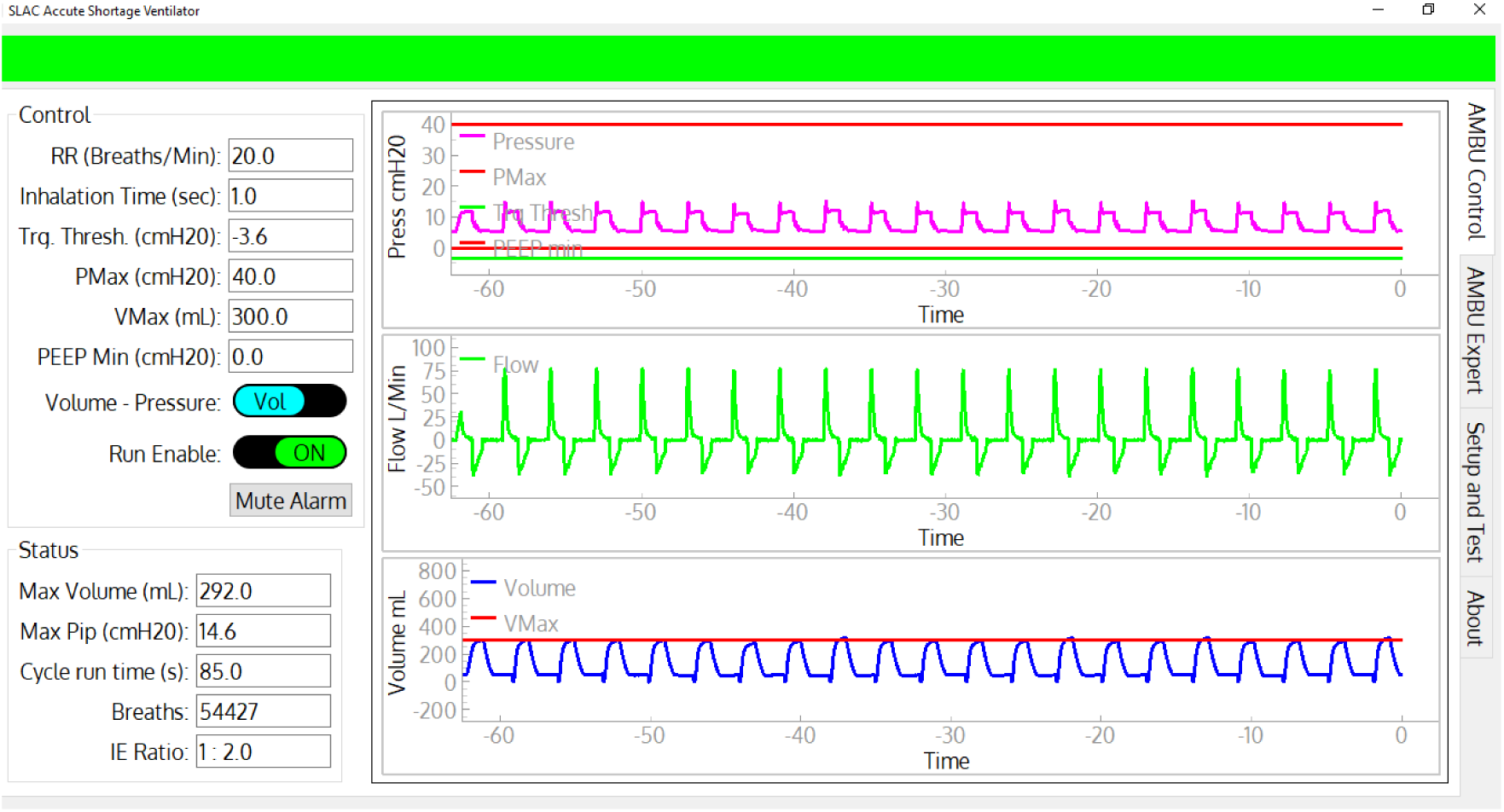
Screenshot of the Control and Monitoring GUI. All primary controllable parameters described above are on the upper left. The time histories, along with the levels for PMax, Trigger Threshold, PEEP Min, and VMax are displayed on the right. The major status values are on the lower left. The green banner on the top indicates no alarm conditions. Given an alarm, it changes to red or amber, flashes, and displays the alarm condition text.

In VC-AC mode, a Volume Maximum (VMax) is set. The flow measured by the spirometer is integrated to calculate the volume, and the bag compression is stopped when VMax is reached. An adaptive algorithm adjusts the precise stop point to accurately approach VMax. In VC-AC mode, the PIP Valve and PMax setting to the microcontroller provide independent limits on the airway pressure.

In PC-AC mode, the target inspiratory pressure is set manually on the PIP Valve. The pressure sensor provides an independent check on the pressure; it halts the bag compression for the current cycle if the PMax parameter is exceeded and it generates a high-priority alarm.

In both modes, the bag is held in its compressed state until the Inspiratory Time is reached. The bag is released, lowering the pressure, and exhalation occurs through the exhaust port of the Patient Valve into an optional PEEP Valve (set manually). The cycle then repeats, triggered either by the RR clock or by a voluntary inhalation.

### 3.3 Controllable Parameters

The fundamental controls for the ASV are:

- Run Enable (On/Off) controls the basic state of the ASV.
- Control Mode (Pressure/Volume) determines whether the ASV is limited by pressure or tidal volume.
- Respiration Rate (RR) in breaths per minute (bpm): This parameter sets the minimal cycle rate for inhalation. Inhalations can occur more frequently by voluntary inhalation when pressure goes below the Trigger Threshold parameter (See below). It is set from the GUI or control panel in increments of 1 bpm.
- Inspiratory Time in seconds: This parameter sets the time for the self inflating bag compression and hold. It is set from the GUI or control panel in increments of 0.1 seconds.
- Peak Inspiratory Pressure (PMax) in cm H_2_O: This parameter is set by manually adjusting the PIP Valve and by setting the PMax parameter in the GUI or control panel. The PIP setting in the software provides an independent alarm if the PIP Valve malfunctions.
- PEEP in cm H_2_O: This parameter is set by manually adjusting the PEEP Valve in the patient circuit.
- PEEP Min in cm H_2_O: This parameter sets a warning threshold if the patient pressure goes below this value.
- Trigger Threshold in cm H_2_O: If the patient pressure goes below this threshold, an inspiration cycle is initiated. This threshold is compared against an absolute pressure gauge measurement, and is not relative to PEEP.
- VMax in ml: This parameter sets the tidal volume limit by stopping the compression of the bag. An adaptive algorithm adjusts the internal microprocessor set-point based on the flow measurement to achieve reasonably precise volume control. This limit is always active, and does not require a different mode of ASV operation.

### 3.4 Monitoring and Alarm System

The ASV monitors status of electrical power and key operational data. It generates medium and high priority alarms annunciated by a sound generator and visual signals on the GUI and local display. A summary of the required alarms is found in Table 2. The visual indications remain on the display until the alarm condition is no longer met. The alarm conditions are:

- Electrical Power Lost (Priority level: High). The 12 V electrical power is lost. The ASV cannot run in this state, but the alarms will be powered by the standby battery.
- PMax (High). The patient-circuit pressure has exceeded the PMax parameter. This indicates there may be a problem with the PIP Valve, the hoses, the endotracheal tube, or the patient. The ASV continues running, but stops the compression of the bag at PMax on each cycle.
- Pressure Low (High). The patient-circuit pressure is low. Possible reasons are the ASV has stopped due to loss of the pneumatic system, or a disconnected patient hose.
- Volume Low (High/Medium). In PC-AC mode this is a high alarm when the volume drops below 250 ml. In VC-AC mode this is a medium alarm when the volume drops below 80% of VMax. The expected volume has not been met, possibly due to disconnected or kinked hose.
- Volume High (medium) In VC-AC mode this is a medium alarm when the pressure exceeds 120% of VMax.
- 9 V Battery Low (Medium). The 9 V standby battery must be replaced to prevent failure of the “Electrical Power Lost” high-priority alarm. However, in the absence of this 9 V battery’s power, the ASV continues running normally.

### 3.5 Mechanical and Pneumatic Subsystems

A self-inflating resuscitator bag is compressed by a pneumatic cylinder inside a simple frame (Figure 1). The frame is made from 18 mm Baltic Birch plywood. The bag is loosely restrained by two plywood stanchions, and is easily replaced for a new patient. The pneumatic cylinder uses a 1-1/16-inch-diameter cylinder with a 4-inch stroke. A 90-mm-diameter disk is attached to the end of the cylinder, which compresses the bag. The bag appears to be resilient enough for a month of steady operation.

The cylinder is controlled by a 3-port 2-position pneumatic valve (PV1) which connects the cylinder to the air supply or to an exhaust control valve. This exhaust valve (PV2) holds air in the cylinder, which gives the possibility of holding the piston down until the Inspiratory Time has been reached.

The ventilator requires a source of compressed air at about 350 kPa (50 psi). This is usually available from a central source in hospitals, but also could be provided by cylinders of compressed air or by air compressors. The ASV consumes about 3 liters per minute (0.1 scfm) at an RR of 20 bpm, thus a standard K-size gas cylinder would support a ventilator for about 30 hours. This is likely about half the rate of oxygen consumption.

The pneumatic circuit is shown in Figure 4. Two manual adjustable flow valves (MV1 and MV2) are used to control the piston compression and return rates. PV1 controls the piston down-stroke and dwell, and PV2 controls the return of the piston. This design allows the piston to stop when the desired volume is reached and maintain pressure in the self-inflating bag for the Inspiratory Time.

**Figure 4:**
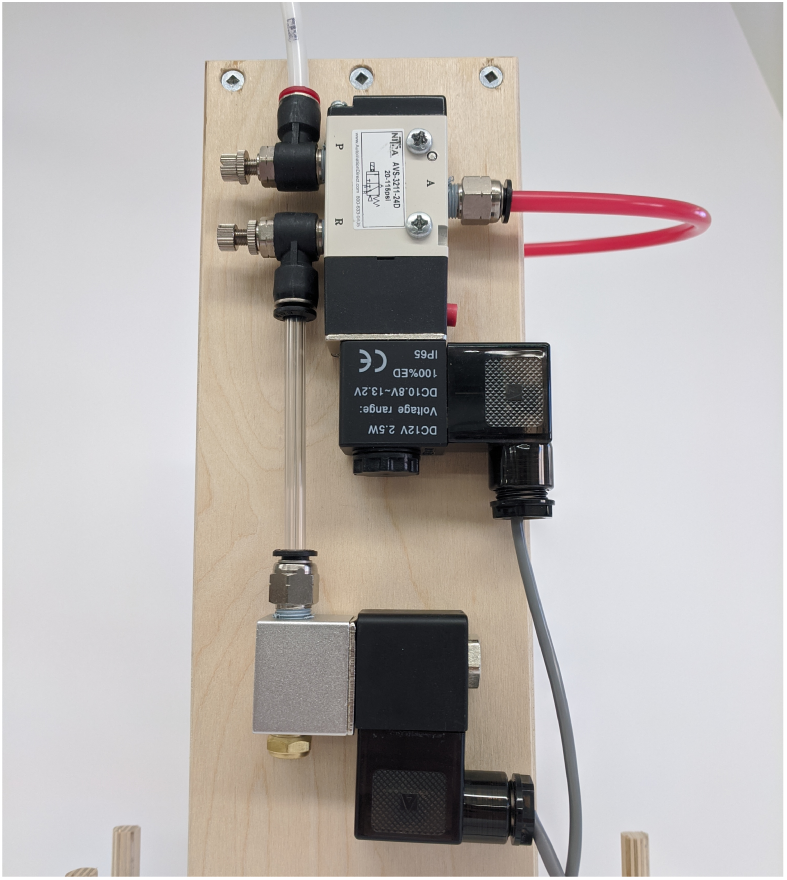
Close-up view of the pneumatic valves. The compressed air input is in the upper left.

### 3.6 Sensors

There are two pressure sensors used to determine the flow and pressure of air in the patient circuit. The sensors are sampled at 100 Hz. The flow measurement is performed by a differential pressure sensor (Superior SP110SM02) with an adjustable range, set to *±* 5 cm H_2_O. Flow is calculated from the pressure drop measured across the spirometer. The patient airway pressure is also measured using a differential pressure sensor (Amphenol DLC-L20D-D4), with one of the two ports connected to the spirometer and the other open to atmospheric pressure. This sensor has a range of *±* 50 cm H_2_O.

The spirometer was initially calibrated against a high accuracy flowmeter [2]. The spirometer flow is proportional to the square root of the differential pressure, and the coefficient is determined from the measurements shown in Figure 5.

**Figure 5:**
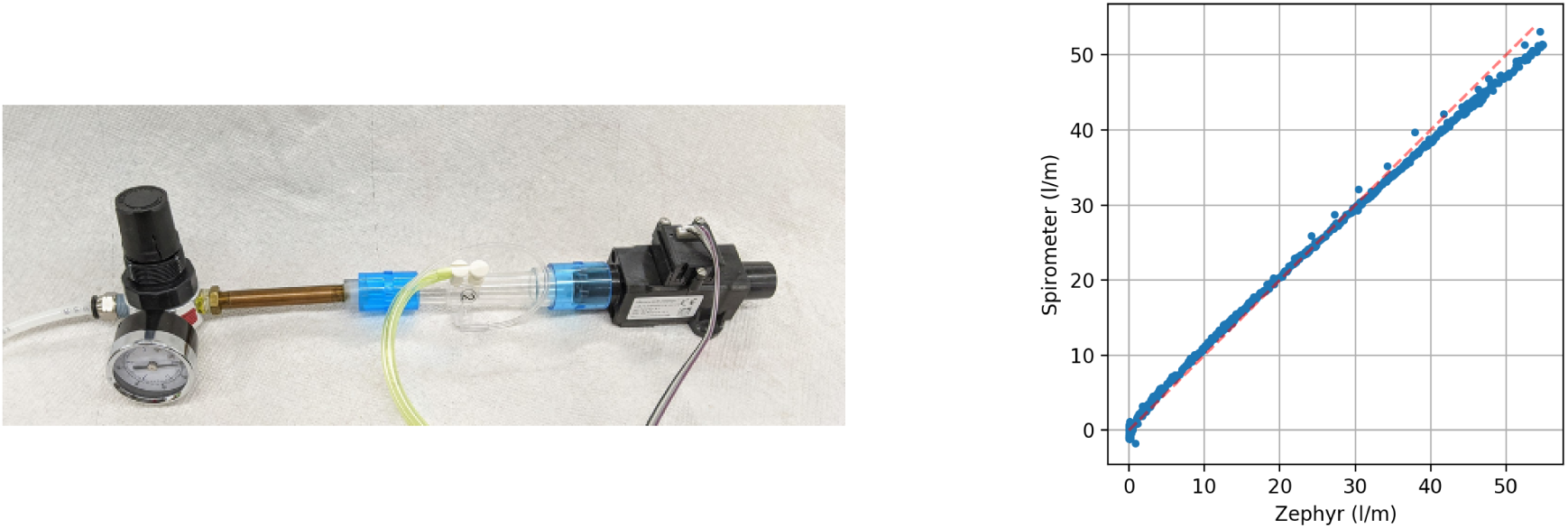
Calibration of the spirometer. (Left) calibration setup showing the spirometer in series with the Honeywell flow meter and a small pressure regulator to sweep the flow. The results are shown in (Right).

### 3.7 Electronics

The primary MCU controlling the operation of the ASV is an Arduino Nano 33 IoT [5]. A second MCU, also an Arduino Nano 33 IoT, runs the integrated display (LCD) and input knob used to display data and parameters and for setting ASV parameters when operating without a computer. Two MCUs allow the separation of critical real-time control of the ASV from the non-critical display and human input. Both MCUs are on a custom circuit board that provides connectors for power input (12 V DC), battery backup (9 V), computer data output (RS232), and connectors to the external pressure sensor box and solenoid valves. Main power is provided to the ASV by a separate medical grade 36-watt AC-to-DC power converter, which avoids introducing line power into the enclosure. Internal backup power is provided via a single 9 V battery, which serves only to maintain the LCD display and alarm capabilities. An external UPS is required for fully-functional uninterruptible power backup to the ASV.

### 3.8 Software

The software consists of three separate components.

1. A real time control component implemented in C++ that runs on the primary MCU. It reads the sensors, operates the valves based on sensor data, time and configuration settings, and sends sensor data via separate serial interfaces to the display controller and, if connected, to the laptop computer. It can also receive configuration commands through the serial connections. Since it is the most critical software component for operation, it has been kept simple for easy review and great care has been taken to make sure communication with the other components cannot block its operation.
2. A display control component, also implemented in C++ that runs on the secondary display control MCU. It receives and displays real-time information, manages a simple configuration menu controlled by an encoder knob, and sends configuration parameter updates to the real time controller.
3. A full GUI implemented in Python that runs on a laptop. It is not required for normal operation but when connected provides the most comprehensive graphical display of system status, sensor data and calculated values. Unlike the built-in display, the GUI provides access to all configuration parameters. The GUI software has been designed to handle disconnects/reconnects gracefully and to clear its state when it detects a new ASV so that a single laptop can be used to control multiple ASVs by connecting to them one at a time. The first tab of the GUI is shown in Figure 3.

The embedded software should be pre-loaded on the MCUs. The GUI software for the external laptop should be distributed on memory sticks or equivalent with the system and requires an existing Windows 10 installation. All software components can be upgraded in the field if necessary.

## 4 Operation of the ASV

A laptop loaded with the GUI executable should be used to verify the functionality of each ASV unit before use. After connection of the services described above and connecting the laptop to the ASV by the supplied serial cable, the GUI should be active. A Setup and Test tab in the GUI guides the user through the setup/checkout procedure.

### 4.1 Setup and Test

The Setup and Test tab uses a sequence of scripts and commands to the primary controller to guide the user through this phase, with the ASV and patient circuit connected to a simple test lung. The protocol ensures that the flow and pressure sensors are properly connected; that the system is reasonably leak tight; and that the PIP Valve vents at its indicated top pressure. Both the VC-AC and PC-AC modes are tested for normal functionality, and the alarm functionality is tested by exceeding a set PMax in PC-AC mode.

### 4.2 Control interface

The pressure flow sensors are used for time history displays, along with a volume computed by integrating the flow. The laptop display is shown in Figure 3. The Respiration Rate (RR) in bpm and the Inspiratory time in seconds are settable. If the patient is not breathing voluntarily, the ASV will operate with the set RR. If the patient begins to inhale before the next scheduled inhalation, as indicated by the patient circuit pressure dropping below the operator-specified Trigger Threshold, a ventilator inhalation cycle will be initiated and the timing cycle reset.

## 5 Performance Testing

The ASV was subjected to increasingly sophisticated performance testing during development and to verify the performance of the final device. Early testing began with simple rubber-bladder test lungs, followed by extensive tests and design feedback using a Michigan Test Lung [3]. The Michigan lung allowed for adjustable compliance and resistance testing. Finally, two rounds of comprehensive tests were performed in a hospital simulation center to determine if the ASV functions correctly as a ventilator and achieves the requirements described in Section 2. A total of 29 scenarios, each with their own set of patient and ventilator variables, were created in collaboration with healthcare experts. The simulated scenarios addressed both healthy and sick lungs, with both passive and active breathing to test the ASV in realistic situations. This section describes the results from these tests to determine if the ASV achieves all the performance requirements described in Section 2.

### 5.1 Test Setup of a High-fidelity Active Servo-Lung Simulator

Final testing was conducted testing using a high-fidelity lung simulator, the ASL–5000 [1], which was made available by the VA Palo Alto Health Care System’s Simulation Center. The ASL (Active Servo Lung) is servo-controlled such that it can replicate with high precision a number of parameter settings for different aspects of patient lung mechanics. These include the “lung model” of lung compliance, inspiratory and expiratory resistances, as well as complex multi-parameter models of the pattern of spontaneous breaths.

Testing was done both with a totally-passive simulated patient (i.e., equivalent of deep sedation and medical drug-induced neuromuscular paralysis) and with the simulated patient spontaneously breathing. Settings for spontaneous breathing used a half-sinusoidal profile of inspiration with the given patient respiratory rate (breaths per minute), maximum inspiratory pressure (−5 or -2 cm H_2_O), inspiratory time (20% of cycle), and expiratory release time (15% of cycle). We did not test with a patient inspiratory pause or patient active expiration. Maximum Inspiratory Pressure is the largest negative pressure generated by the patient’s own breathing. Since ventilators are commonly set to trigger a mechanical breath at a patient breath of between -2 and -5 cm H_2_O, we used these settings to test the ability of the ASV to trigger correctly.

The ASL–5000 was equipped with a thorax, head, and neck mannequin which allowed the insertion of an endotracheal tube (7.0 – 8.0 mm internal diameter) to replicate the situation and tube resistance of a patient requiring mechanical support or ventilation.

### 5.2 Test Scenarios

Each test used a specific lung model and inspiratory pattern, and lasted at least twenty breaths in steady-state. This condition was reached quickly for the passive simulated patient, but took longer when the simulated patient was breathing spontaneously. Data were sampled at 64 Hz by the ASL–5000. The variables used in this analysis were: flow, airway pressure, lung pressure, muscle pressure, and volume. Multiple waveforms were visible in real-time for inspection, and the waveforms and computed results stored automatically by the simulator.

Tests with the ASL–5000 covered different lung and breathing models (see Table 3): normal and ARDS with both passive and spontaneous lungs. The ARDS tests were performed with three different severities by setting the lung compliance and resistance. Spontaneous ventilation varied for respiratory rate (10 – 30 bpm) and maximum inspiratory pressure (−2 or -5 cm H_2_O). For each lung model, several tests were done with different setting of the ASV. Table 4 shows the different modes of the ventilator and ranges for the different settings.

**Table 3:** ASL–5000 settings for different lung models tested.

| Model | Lung Compliance<br>[ml/(cm H<sub>2</sub>O)] | Inspiratory Resistance<br>[ml/(s cm H<sub>2</sub>O)] | Expiratory Resistance<br>[ml/(s cm H<sub>2</sub>O)] |
| --- | --- | --- | --- |
| Normal | 50 | 6 | 6 |
| ARDS 1 | 30 | 6 | 6 |
| ARDS 2 | 30 | 11 | 16 |
| ARDS 3 | 10 | 20 | 20 |

**Table 4:** Overview of tested ASV settings for the volume- and pressure-control ventilator modes. The selected ranges were determined by the EURS design specifications and further informed by medical expertise.

| Ventilator Mode | RR [bpm] | Set Tidal Volume [ml] | Peak Insp. Pressure [cm H<sub>2</sub>O] | Insp. Time [s] | I:E | Trigger Threshold Pressure [cm H<sub>2</sub>O] | PEEP [cm H<sub>2</sub>O] |
| --- | --- | --- | --- | --- | --- | --- | --- |
| Volume Control/<br>Assist Control | 10–30 | 250–600 | 15–35 | 0.6–1.5 | 1:1–1:4 | -2– -5 | 5–20 |
| Pressure Control/<br>Assist Control | 20–40 | NA | 25–35 | 1.0 | 1:1–1:2 | NA | 5–20 |

The data for two of the tested scenarios are shown in Figure 6. Here, the same ventilator settings (in VC-AC mode) are used for two different lung models, with the only exception being the PEEP value, which acts as a pressure offset. For both scenarios three example breaths are shown for the steady-state regime. A single breath starts with the flow (blue curve) rapidly rising, which is caused by the piston moving down and squeezing the self-inflating bag. The rate the piston descends is adjustable by the needle valve input to the pneumatic system, which is set here to about half a second, until it hits the VMax set-point and is then held in place. During this time the pressures (green and red curves) stay high but the flow goes to zero. This is what a patient would experience as an inspiratory pause and increases the time oxygen can be absorbed in the lungs.

**Figure 6:**
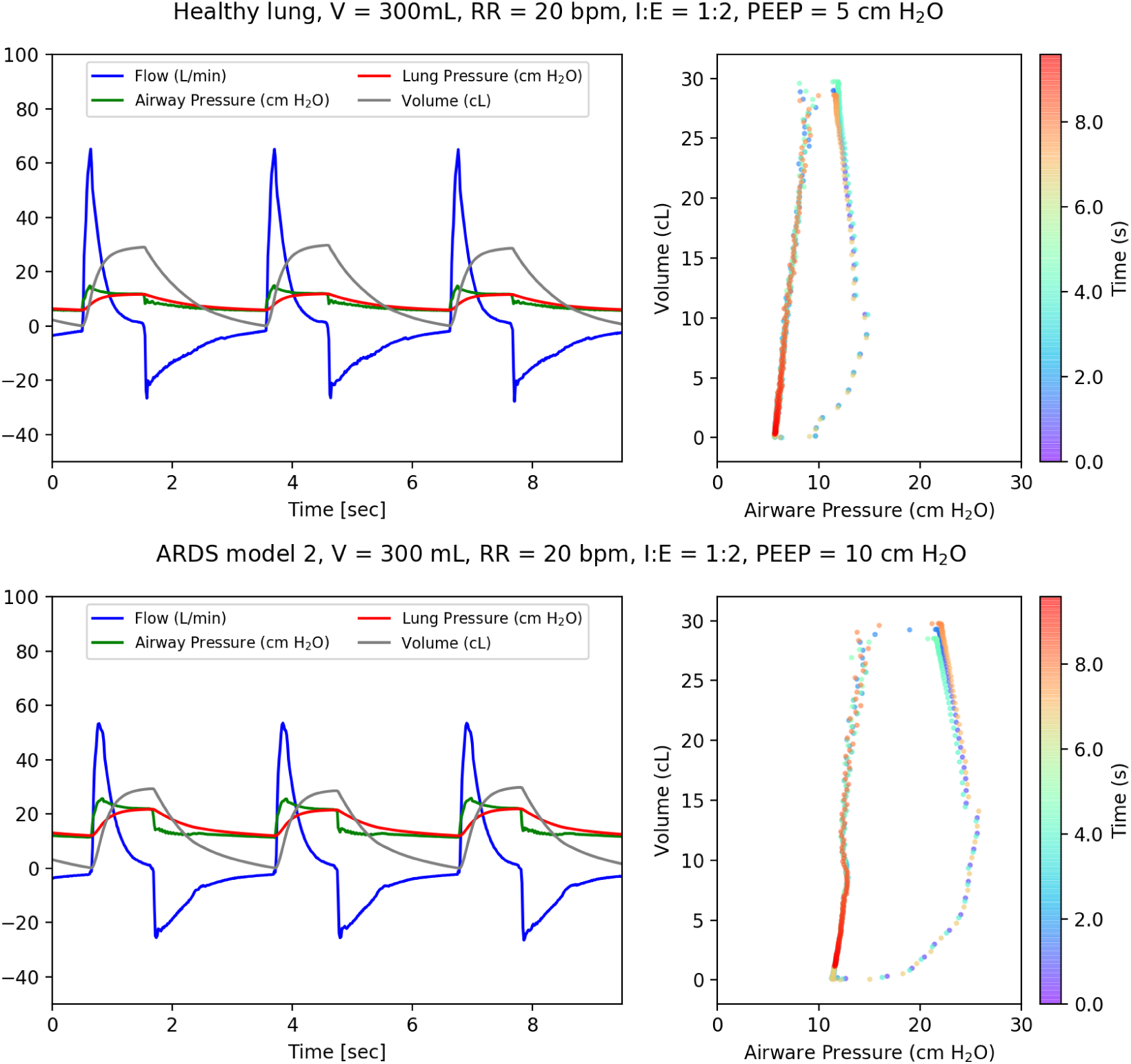
Example waveforms from two of the tested scenarios. Top left: ventilator connected to the simulator with a healthy lung model showing the flow (blue), pressures (red and green) and volume (gray). Top right: for the healthy lung data the pressure-volume curve over time (in color). Bottom left: ventilator connected to the simulator with a ARDS 2 lung model showing the flow (blue), pressures (red and green) and volume (gray). Bottom right: for the ARDS lung data the pressure-volume curve over time (in color). In both scenarios the ASV reaches the set goals, but as expected the pressure-volume curves show that a higher pressure is needed to reach the same volume in the ARDS lung than in the healthy lung.

Once the inspiratory time ends the piston moves upwards and the simulated patient exhales. The start of exhalation is shown by the volume (gray curve) decreasing after it has reached its maximum and by the flow reversing. The difference between the measured lung and airway pressure is due to the resistance of the lungs in the simulated patient. On the right side of the figure the pressure-volume curves are shown for both test scenarios. It is apparent that for a sick lung a higher pressure is needed to reach the same volume of air in comparison to a healthy lung. These curves show the ability of the ASV to deliver a set tidal volume of 300 mL in two different lung scenarios by utilizing increased pressure when required for a patient with ARDS.

### 5.3 Test-Data Analysis

The time history of the flow, pressure and volume are analyzed to get performance parameters, including breaths per minute, delivered tidal volume, and peak inspiratory pressure. A minimum of twenty breaths are used for the analysis. For each breath the following parameters (derived values) are determined: minimum pressure (typically, the set PEEP), maximum pressure (PIP), respiratory rate (bpm), the ratio of inspiratory to expiratory time (I:E), and the integral of the flow (tidal volume). For every derived parameter, we report the mean and standard deviation.

### 5.4 Demonstration of Performance Requirements

To demonstrate that the ASV achieves the tidal volume range requirement of 350 to 450 ml, or preferably 250 to 600 ml (EURS-59), we tested the ventilator in both VC-AC (Volume Control - Assist Control) and PC-AC (Pressure Control - Assist Control). Figure 7 shows the delivered tidal volume versus the set tidal volume for the ventilator in VC-AC mode while connected to the ALS-5000 normal lung model (refer to Table 3 lung model details). For the five measurements, over a total range of 250 to 600 ml, the delivered tidal volume was within the 15% required tolerance.

**Figure 7:**
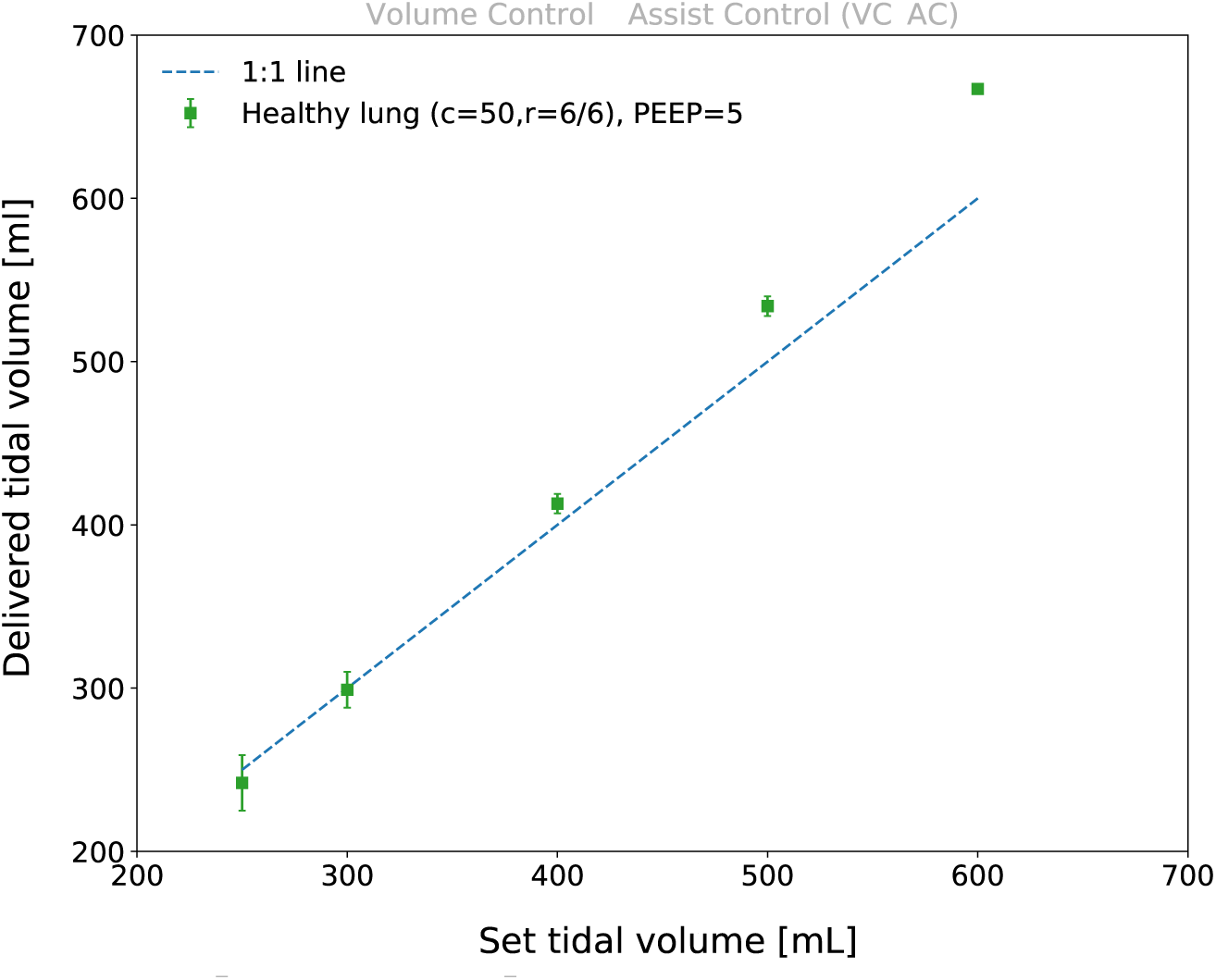
Delivered tidal volumes with the healthy lung model as a function of set tidal volume on the ASV in VC-AC mode connected to a passive lung. The dashed line is to guide the eye in seeing departures from one-to-one linearity. The required delivered tidal volume is 350 to 450 ml (EURS–59). As defined in the text, *c* is the compliance and *r* is the resistance (inspiratory / expiratory).

To explore the delivered tidal volume as a function of measured PIP, several tests were done in PC-AC mode. Figure 8 shows the delivered tidal volume and measured PIP for five different ventilator settings on four different lung models (Table 3). These measurements are all taken with the ASV in PC–AC mode and show the maximum tidal volume the ASV can deliver. A higher PIP is required to reach the maximum tidal volume for more severe ARDS models. For the healthy lung, another measurement was made at a lower PIP, resulting in a lower delivered tidal volume. For the ARDS configurations, the PEEP Valve was manually set to 10 cm H_2_O. PEEP values of 10, 15, or 20 cm H_2_O are consistent with what is commonly utilized for patients with significant ARDS. The respiratory rate was kept at 20 bpm, the I:E ratio at 1:2, and the PIP Valve at 35 cm H_2_O. These results show that the ASV in VC-AC or PC-AC modes achieves the required tidal volume range (EURS-59) for all scenarios except for the most extreme ARDS 3 model, where it achieved only 259 ml.

**Figure 8:**
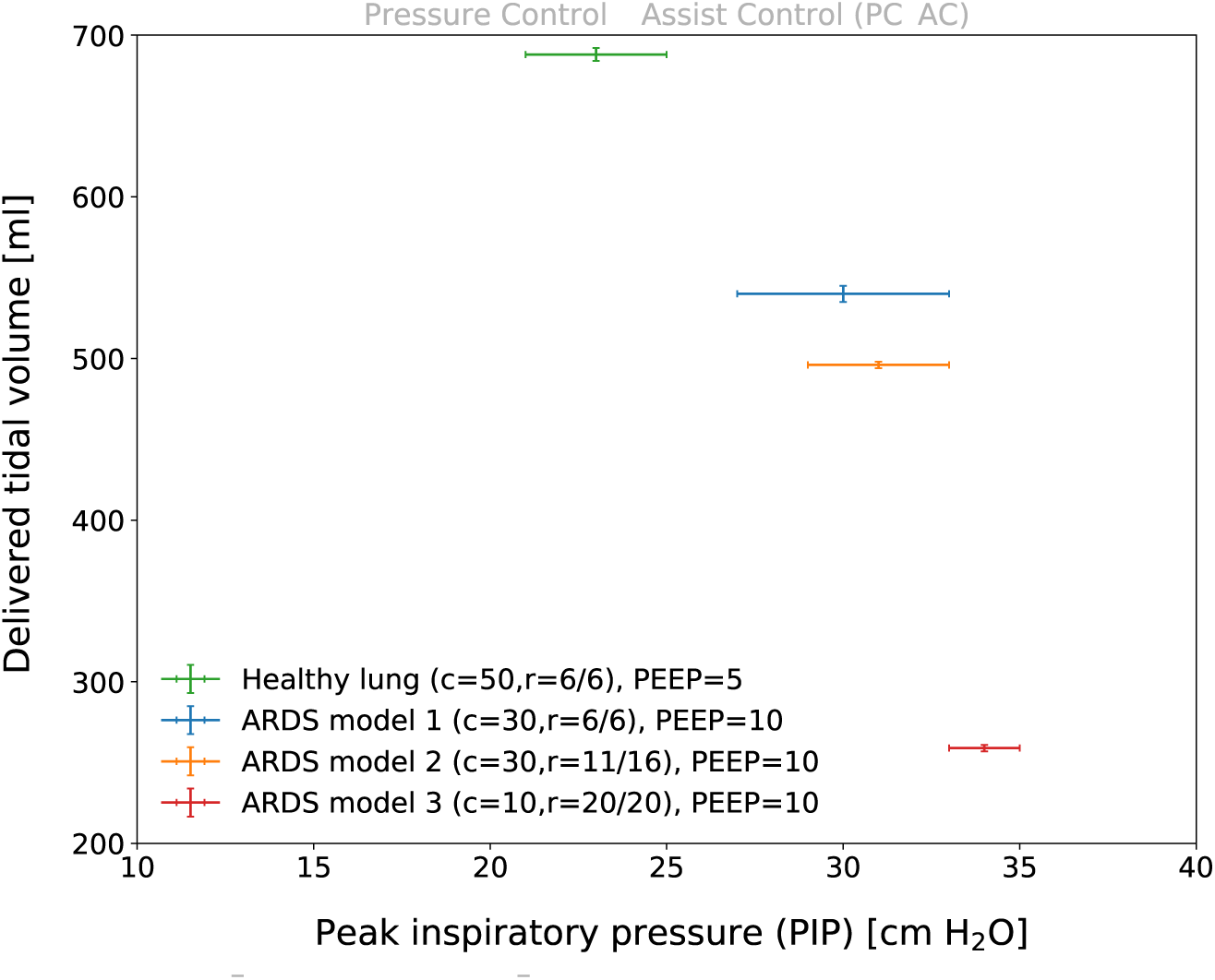
Delivered tidal volume as a function of set peak inspiratory pressure (PIP) with the ASV in pressure-control mode connected to the ASL–5000 set to passive-patient mode, where *c* is the compliance and *r* is the resistance (inspiratory / expiratory).

The ASV was tested on a healthy lung in VC-AC mode with respiratory rates between 10 and 30 bpm. It achieved the required respiratory rate range of 10 to 30 bpm (EURS-57) within the tolerance of 2 bpm.

Table 5 shows the measurements of the inspiratory to expiratory time (I:E) for different inspiratory times. All four measurements are acquired with the ASV in VC-AC mode with a set tidal volume of 400 ml, a PEEP value of 5 cm H_2_O and a PIP value of 35 cm H_2_O. As all measurements are done with the same respiratory rate (RR), only the inspiratory time is changed to change the I:E ratio. For a respiratory rate of 20 bpm the breath cycle time is a total of three seconds. The piston speed was set with the needle valve to achieve a full downward stroke in 0.5 seconds; the rest of the inspiratory time is thus used as an inspiratory pause. During the inspiratory pause, the pressure is kept constant so that the lungs stay fully open before the piston retracts and the patient exhales. As shown, the ASV is able to achieve the required range of the I:E ratio of 1:1 to 1:4 (EURS-56).

**Table 5:**
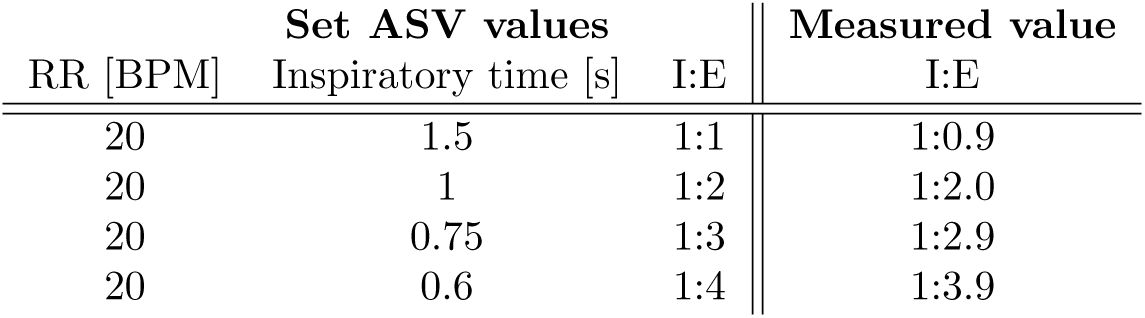
Set and measured inspiratory to expiratory time (I:E) for the ASV in VC-AC mode connected to a passive lung. The measured uncertainty on the I:E ratio is less than the rounding error. The required range of the I:E ratio is 1:1 to 1:4 (EURS-56).

The maximum peak inspiratory pressure (PIP) is controlled by the PIP Valve that functions as a “pop-off” valve as described in Section 3. Therefore, as expected, the measured PIP values for all 29 tested scenarios stayed below the required 40 cm H_2_O (EURS-256). The highest reported PIP value was 34 *±* 2 cm H_2_O.

The ASV provides spontaneous breaths in compliance with EURS-43, which was demonstrated by using the ASL–5000 in active mode. Figure 9 shows the waveforms for the lung simulator requesting 30 bpm with an inspiratory pressure of -5 cm H_2_O coupled to the ASV in VC-AC mode. The ASV settings were 20 bpm, 400 mL tidal volume, PEEP of 5 cm H_2_O, and a Trigger Threshold of 3 cm H_2_O, where the ASV Trigger Threshold pressure is set to -2 cm H_2_O with respect to the PEEP. A spontaneous simulator breath is started by decreasing the inspiratory “muscle pressure” (red line). When the airway pressure drops below the Trigger Threshold the ASV starts a breath, as indicated by a sharp (positive) rise in the flow (blue line). If the ASV RR is small compared to the spontaneous breathing rate then the breathing cycles can be missed because the airway pressure does not fall below Trigger Threshold (as seen around 17s time). If the Trigger Threshold is not met in the time window set by RR, a breath will be provided by the ASV at the appropriate time.

**Figure 9:**
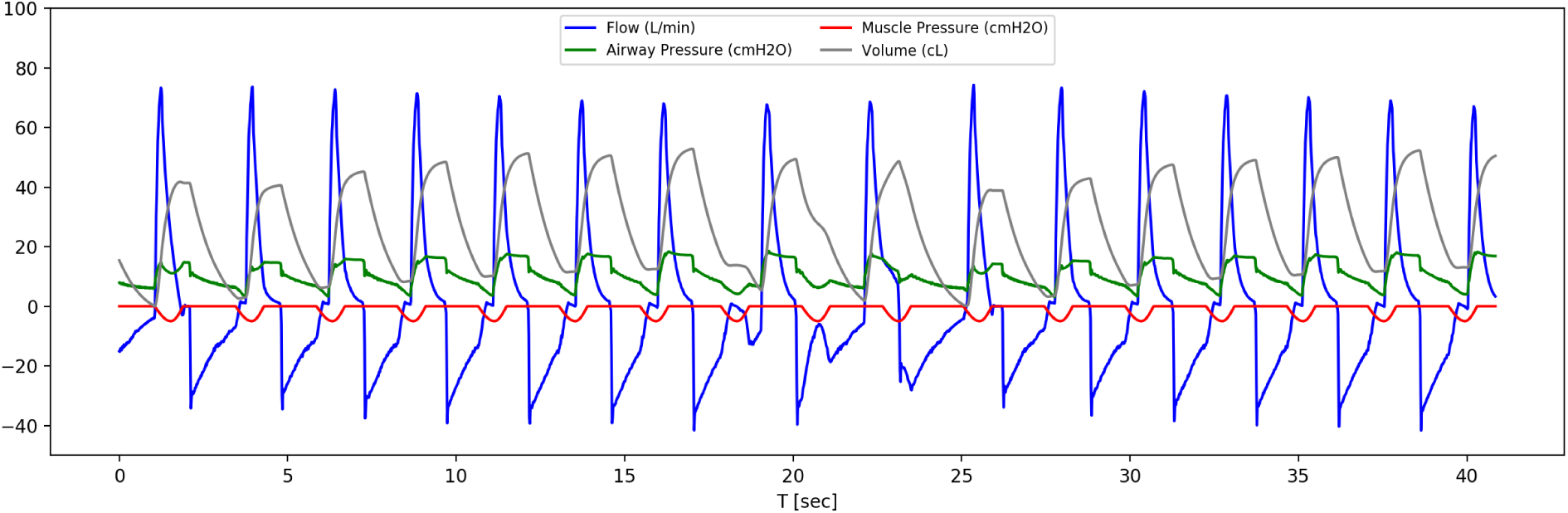
Example waveforms of the ASV set to 20 bpm triggering on the spontaneous breathing of the lung simulator set to 30 bpm. The muscle pressure (red line) shows when the spontaneous breaths are requested by the lung simulator, while the start of the positively increasing flow (blue line) shows when the ASV delivers a breath. The ASV correctly triggers on most breaths, but because the exhalation time setting is too short at 30 bpm, the pressure does not reliably fall below the Trigger Threshold before the simulated patient requests the next breath. With these settings about one in ten spontaneous breaths is missed, falling back to the ventilator timing.

Breathing through a disabled ASV was tested by an author breathing through a mouthpiece connected to the spirometer port. The ventilator was set to only record the data, and Figure 10 shows the waveforms for this test. As expected, breathing was possible as there is an inlet valve on the self-inflating bag. In any case, this causes a high priority alarm.

**Figure 10:**
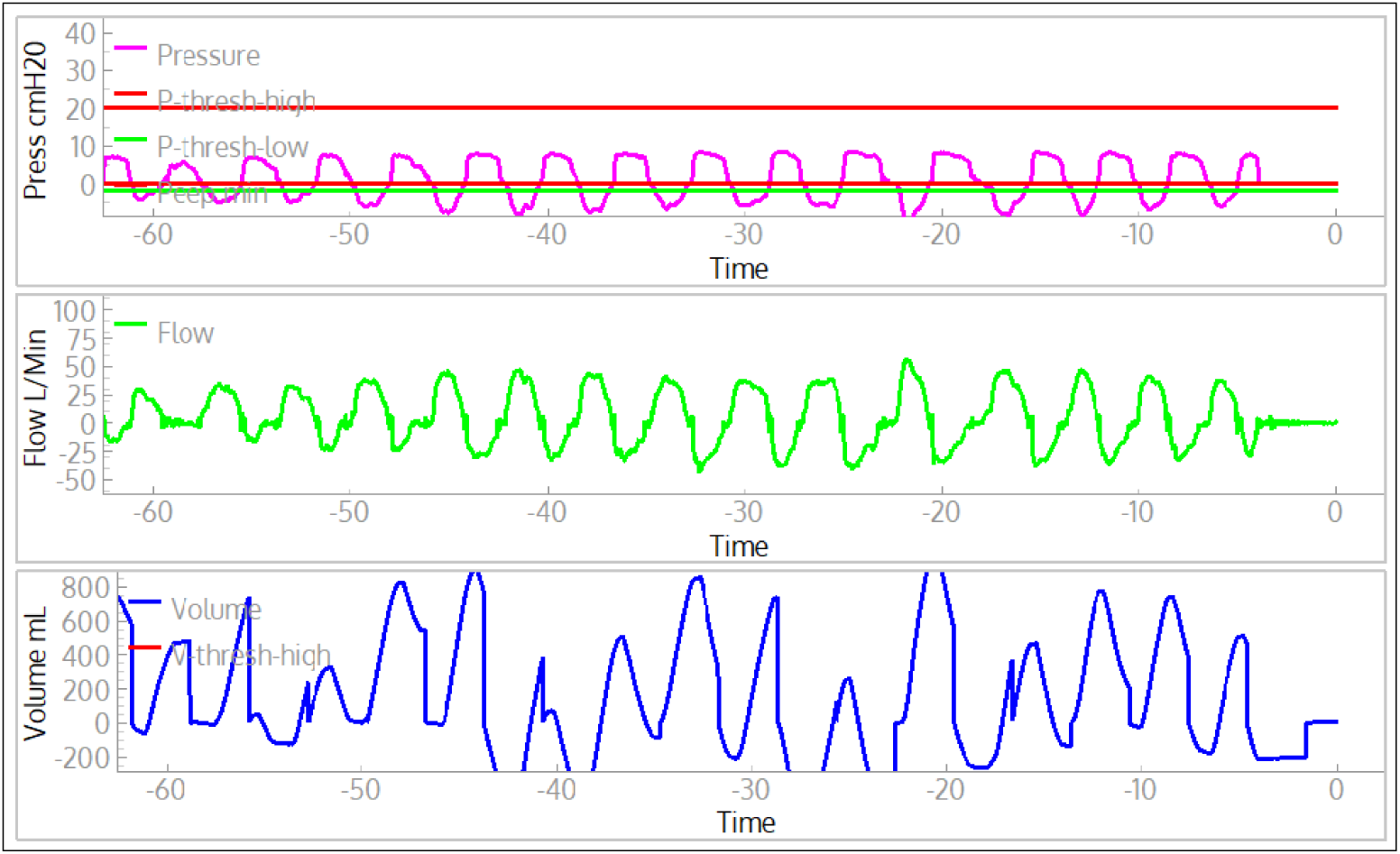
Example waveforms of one of the authors spontaneous breathing with a ventilator attached, but only in monitoring mode. This shows that a normal person can breathe voluntarily even if the ventilator is completely turned off; the valve on the self-inflating bag allows for this.

## 6 Summary and Discussion

In this paper we described the design, implementation and testing of a low-cost easy-to-manufacture Acute Shortage Ventilator, or ASV, for potential use by medical professionals during shortages of standard equipment, motivated by the COVID-19 pandemic. Our device consists of a patient circuit based primarily on a readily-available self-inflating resuscitator bag and other standard ICU parts, together with a pneumatic plunger system to automatically compress the bag based on adjustable parameters. The patient circuit consists of only standard hospital parts to help medical professionals have sufficient confidence in the device should they require its use for treating patients in their communities. Additionally, by selecting primarily non-medical components for the compression system, as well as related sensors and control elements, we sought a design that as much as possible took advantage of items in industrial supply chains. Pressure and flow are measured by modern differential pressure sensors that are read by microcontrollers and permit volume- or pressure-control assisted breathing. The ASV can be operated stand-alone with the integrated display and controls. An optional Windows-10 computer and Graphical User Interface provides full time-history plotting.

The development of the ASV was carried out through a partnership of SLAC National Accelerator Laboratory scientists and technical staff, and medical professionals from three Bay Area medical centers, the Stanford University School of Medicine, Santa Clara Valley Medical Center, and the VA Palo Alto Health Care System. Technical development and testing began with simple rubber-bladder test lungs, followed by extensive tests and design feedback based on tests with a Michigan Test Lung, and finally, two rounds of performance tests with the ASL–5000. The final round of tests with the ASL–5000, a high-fidelity test lung simulator system, demonstrated that the ASV meets all requirements defined by the EURS specifications. We welcome contact from parties interested in producing and distributing this device. The intellectual property of the ASV will be held by Stanford University and made available to vendors and manufacturers via no-cost licenses. The combination of high-performance, easy manufacturability, and low-cost of the ASV may make it an effective tool in combating the COVID-19 pandemic.

## Data Availability

All data used to determine the performance of the ventilator is freely available by contacting the corresponding author

https://www.slac-asv.net/

## 7 Acknowledgements

We acknowledge important suggestions by K. Skarpaas VIII and technical support from R. Conley, both of SLAC National Accelerator Laboratory. We thank Aaron Roodman of SLAC National Accelerator Laboratory for early suggestions, especially about the use of the spirometer for flow measurement. Paul Barraza, a respiratory therapist at Santa Clara Valley Medical Center, provided essential encouragement and invaluable contributions to our ventilator project. Because of his clinical experience with critically ill, ventilated ICU patients, he was able to help the team better understand the multiple components of a mechanical ventilator, and the issues involved in initiating mechanical ventilation, maintaining successful mechanical ventilation, and dealing with complications. We acknowledge Kyle Halkola for his extremely helpful advice and explanations of ventilator technology and the methods of ventilating patients. We also acknowledge Cynthia Shum and Devin Stine of the Simulation Center at VA Palo Alto Health Care System. We thank J. Aalbers of Stockholm University for his help on creating the data analysis pipeline.

Research was supported by the DOE Office of Science through the National Virtual Biotechnology Laboratory, a consortium of DOE national laboratories focused on response to COVID-19, with funding provided by the Coronavirus CARES Act, and through Stanford University discretionary funds allocated to SLAC National Accelerator Laboratory. SLAC National Accelerator Laboratory is supported by the U.S. Department of Energy, Office of Science under Contract No. DE-AC02-76SF00515.

## Notes

### Competing Interest Statement

The authors have declared no competing interest.

### Author Declarations

This research was conducted by slac (legal https://legal.slac.stanford.edu/), this research did not involve humans and no approval nor exemption is necessary.

### Summary of Updates

Updated funding acknowledgments to explicitly name the CARES act

